# Developing clinician-centred design principles for Genomic Test Reports in Pakistan

**DOI:** 10.1101/2023.12.03.23299329

**Authors:** Momal Agha, Faisal F Khan

## Abstract

Despite rapid technological progress being made in genomics, a growing disparity is emerging between healthcare in developed and developing countries. This *genomic divide* can be partly explained by the scarcity of available genomics workforce and in some parts by limited genomic literacy of healthcare professionals that reportedly deters them from proposing genomic testing in a clinical setting. This study aims to study this gap in a local context and learn how we can reduce this *genomic divide* by developing a user-centred design of genomic test reports in Pakistan. The user being the clinician in this study.

We selected two commonly used genomic reports which varied in language, content, and layout. Report A was a one-page genomic report from the Laboratory for Molecular Medicine at Partner’s Healthcare. Report B was a report with multiple pages of information from FoundationOneCDx. We employed a qualitative descriptive study design, including a survey of trainees, non-specialists, and specialists. The parameters recorded were: subjective comprehension, overall visual impression, level of difficulty of the language, and communication efficacy depending on the reports’ graphical representation, along with actionability and degree of reliability.

A total of 49 medical professionals across 11 institutes in Pakistan participated in the survey. Based on the answers and suggestions provided by the participants, we extracted 11 recommendations and broadly grouped them into four categories, i.e. language, content, layout and reliability.

Our findings highlights key areas that need to be taken into consideration when designing impactful genomic reports for clinicians in Pakistan. This incudes accessible and appropriate language, adequate content and a non-overwhelming and friendly layout as well as an emphasis on establishing reliability and actionability of what the clinician finds in the report. This can be instrumental in helping us improve the adoption of genomic testing in clinics around Pakistan, and potentially in other similar contexts.

## Introduction

Traditional healthcare has undergone a significant transformation since 2003 when the leaders of the Human Genome Project announced that they had successfully sequenced the 20,000, or so, genes responsible for the genetic blueprint of the modern human species (Heggie, 2019). Over the years genomics has greatly enhanced healthcare outcomes such as in early diagnosis and the treatment of diseases before they progress towards severity (Khoury, Burke, and Thomson, 2009). Genetic and genomic testing services for cancer patients as well as prenatal or newborn screening have transformed healthcare approaches from a general one-size-fits-all approach to a more personalized one, and are already making a considerable difference in clinical care (Riaz *et al*., 2019). It is therefore no surprise that Precision Medicine, an emerging area that considers the ‘genes, environment as well as the lifestyle’ of the patient, is growing fast and is expected to expand even further in its applications especially with the steps reduction in sequencing costs of advanced genomic testing platforms which enable whole-genome sequencing and whole transcriptome sequencing.

However, most progress in genomics and public health has occurred in developed economies of the world, and this is leading to a *genomic divide* emerging between healthcare in developed and developing countries (Sirisena *et al*., 2016). Apart from lack of access to the latest equipment and their often proprietary reagents, one of the reasons for this divide is also a shortage of a professionally trained genomics workforce as well as a limited awareness of genomics amongst clinicians in countries like Pakistan (Ashfaq *et al*., 2013, Barakzai *et al*., 2022). The lack of a specialization or any certification being offered by the College of Physicians and Surgeons of Pakistan (CPSP) in genomics or even genetic counselling is exacerbating the problem. Despite these shortcomings, there have been some developments in genetic screening as well as some genomics initiatives such as ‘The Congenital Hypothyroidism Screening Programme’ by the Aga Khan University Hospital (Afroze, Humayun and Qadir, 2008) and ‘Punjab Thalassemia Prevention Programme’ launched in 2012. Moreover, the recently launched Pakistani Society of Medical Genetics (Furqan, 2021) has a goal to spread awareness regarding genetic disorders and testing along with helping the community through telemedicine and is aiming to accelerate the progress towards the adoption of personalized medicine in Pakistan.

Another barrier to adoption is the real or perceived lack of accessibility and actionability of genomic test reports when they reach the clinic. Clinicians have been reported to find it challenging to understand and therefore communicate the genomic test results without prior training (Brett *et al*., 2022). To be able to undertake a clinical decision and also communicate the results of a genomic test to the patient, the clinician has to able to interpret the results as thoroughly and confidently as possible (Recchia *et al*., 2020). For this to happen, we need to pay attention to a very crucial yet overlooked aspect, which is the design and layout of the genomic reports that can aid both specialists and non-specialists in the understanding of results.

According to research, even medical professionals face difficulties in understanding genetic and genomic test results (Sandhaus *et al*., 2001; McGovern, Benach and Zinberg, 2003) and many have realized the importance of clearer and simpler reports with regards to differences amongst them with regards to genomic literacy (Haga *et al*., 2014; Ostergren *et al*., 2015; Joseph *et al*., 2017).

At the time of interpretation of genomic results, there are numerous technical details that need to be understood before making a clinical decision including terms such as the different types of variants, the coverage or depth of sequencing, the different sources of error and understanding what it really means to be looking at a variant of significance (VUS). These challenges are multiplied in cases where more than one gene underlie a particular disease or in many cases when not much is known about the variants in a particular population (Farmer *et al*., 2020).

Such problems have long been recognized by leading North American and European bodies such as the American College of Medical Genetics and Genomics (ACMG) (Richards *et al*., 2015, the European Society of Human Genetics (ESHG) (Matthijs *et al*., 2016), and the Association for Clinical Genetic Science (ACGS) (Cresswell *et al*., 2020), which have published guidelines on the reporting of genomic tests for their audiences. These professional guidelines touch upon technical details and testing methods primarily relating to laboratory reports that are important for establishing and accrediting a genomic testing facility. Moreover, these guidelines have to continuously revise as laboratories adapt to technological advances.

With the technical guidelines relating to the content and details of the report covered by experts in genomics, there still remains a need to understand the design elements including layout, language and length, to be able to produce reports that are accessible to not just patients and their families, but more importantly their doctors. A lack of understanding by patients has been reported to exhibit lower engagement and satisfaction with their treatment (Elder and Barney, 2012), show poor compliance with the recommended interventions as well as undergo stress and anxiety due to overall dissatisfaction (Dang *et al*., 2013; Linn *et al*., 2013). Well thought out reports especially those designed with a user-centric approach that is relevant to local practice, outside the developed world will certainly be helpful. TThere is a growing demand for plain text genomic reports with the increase in adoption of consumer genomics products used to answer questions around wellbeing, ancestry and longevity. Reports produced by direct-to-consumer companies utilise more colours and graphics to differentiate sections when compared to other types of reports. It is possible that these private companies have conducted studies with their users which reveal a preference for such graphics, but this information is not publicly available (Shaer et al., 2015). Finally, a well-designed report may ultimately help in increasing the adoption of genomic testing and challenging the potential disparities or the genomic divide that is increasing otherwise.

In this survey we set out to study the understanding and perception of two physician prescribed genomic test reports (one by Partners Healthcare and another by FoundationOne) amongst medical professionals in Pakistan. We specifically ask them about how they compare the two reports with regards to ease of understanding, overall length, language, actionability and perceived reliability. We then summarise our findings into four categories of recommendations which we hope will help will help in bringing down barriers (Riaz *et al*., 2019) and catalyzing the adoption of genomic testing in clinical practice across Pakistan.

## Methodology

### Selection of report templates

We undertook a literature and market review of existing genomic reports and selected two contrasting reports for our study. The two physician-prescribed genomic report templates, one by the Laboratory for Molecular Medicine at Partners Healthcare (referred to as report A) and other by FoundationOne (referred to as report B) that differed in terms of language, layout, and content.

The reports were chosen based on the following differences in order to prompt comments from specialists and non-specialists alike. Report A was a one-page sample genomic report of a patient with suspected hypertrophic cardiomyopathy, containing concise and relevant details pertaining to the medical diagnosis. Report B was a detailed sample report of a breast carcinoma specimen with the confirmed findings mentioned. The first page of report B had the detected genomic results paired with an FDA-approved therapeutic option, while the rest of the pages consisted of all the details related to the genomic and biomarker findings, along with the technical details about the DNA sequencing procedure.

With these two templates in place, we designed a questionnaire to be filled in, either manually or online, rating the different aspects of both reports.

#### Questionnaire Survey tool (Formative Evaluation)

A total of 49 participants completed the questionnaire form after atleast five minutes of deliberation on both the report templates each. With each of the 11 questions, free text fields were used to give the participants the freedom to comment under the following categories: first visual impressions, ease of understanding, length of the reports, trust in the result, the possibility of action to be taken and report preferences with reasons along with any recommendations based on their expertise. A six-point Likert scale was used to rate the difficulty of the scientific language used in both reports.

Data analysis from the survey forms was performed using a qualitative descriptive approach. All comments under their respective questions were recorded and displayed as frequency tables while the quantitative data was displayed in graphical form. The recommendations and general input given by the doctors were then classified into four themes: communication style, the content of the report, general layout and trustworthiness of the report.

## Results

### Demographics of the Study Participants

Out of the 49 recruited participants, 20.2% were specialists (4.1% Assistant Professors and Registrars each and 2% Senior Registrars, Demonstrator, Lecturer, Senior Lecturer, Consultants, and Head of Department each) while the dominant majority (79.6%) were non-specialists that predominantly included Trainee Medical Officers who are undergoing their 4-year post-graduate training (71.4% Trainee medical officers, 4.1% MPhil students and 4.1% Medical Officers each) as shown in Figure 1.

**Figure 1.**
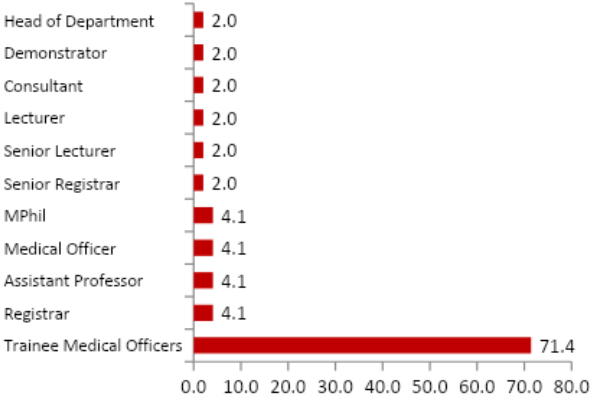
Designation of the participants: 71.4% of the 49 doctors were Trainee Medical Officers, 4.1% were each of Assistant Professors, MPhil students, and Registrars and 2% were each of Senior Registrars, Medical Officers, Demonstrator, Lecturers, Senior Lecturer, Consultants and Heads of Department.

The respondents belonged to 11 different medical institutions with RCD, HMC and RMI in Peshawar dominating the group (Figure 2). The participants come from 12 various specialties including 18.4% from Oral and Maxillofacial Surgery, 14.3% from Prosthodontics and 12.2% from Periodontics (Figures 3).

**Figure 2.**
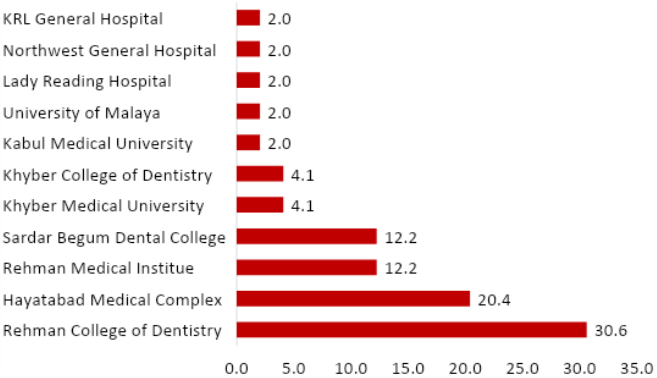
Institutions of the participants: About 30.6% of the participants were from Rehman College of Dentistry, 20.4% were from Hayatabad Medical Complex, 12.2% were from Rehman Medical Institute and Sardar Begum Dental College, 4.1% were Khyber Medical University and Khyber College of Dentistry, whereas the remaining 2% were from Kabul Medical University, University of Malaya, Lady Reading Hospital, KRL General Hospital and Northwest General Hospital.

**Figure 3.**
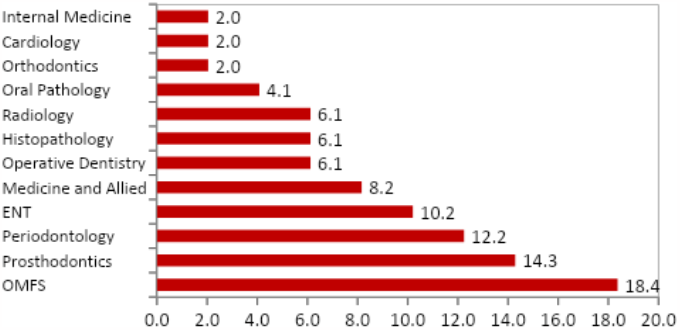
Specialty: The participants in the survey came from 12 different specialties: 18.4% were from Oral and Maxillofacial Surgery, 14.3% were from Prosthodontics and 12.2% were from Periodontics.

### First Visual Impressions of Reports A and B

Participants were asked to describe their first visual impressions regarding reports A and B. Frequent words generated from each report when translated into frequency tables revealed that 38.8% of the medical professionals used ‘concise’ for Report A, 19.4% mentioned ‘easy to interpret’ and 7.5% considered Report A to be ‘well organized’ (Figure 4a). Regarding Report B, 33.8% of doctors found it ‘lengthy’, 32.3% stated that it was overly ‘detailed’ and 12.3% found it ‘exhaustive’.

**Figure 4.**
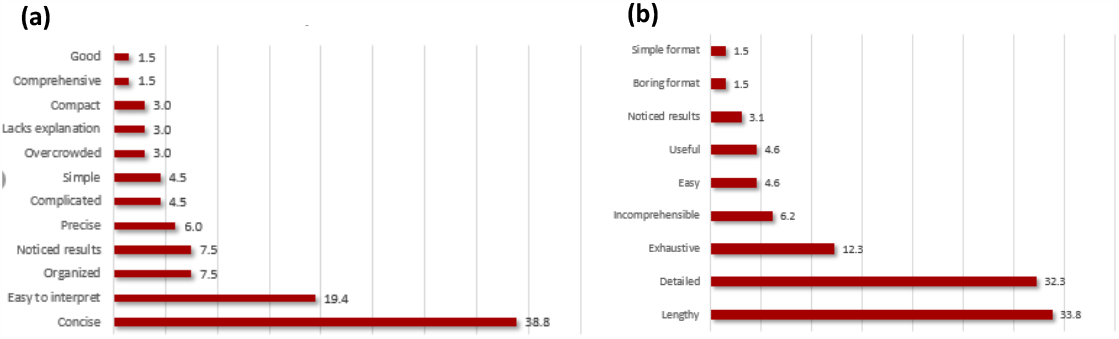
First visual impressions of reports A and B: **(a)** Respondents that described report A to be ‘concise’ were 38.8%, 19.4% answered it was ‘easy to interpret’, 7.5% said they were ‘organized’ and they could ‘notice the results’ straightaway. 6.0% found it ‘precise’, 4.5% said report A was ‘simple’ and ‘complicated’’. 3.0% found it overcrowded’, ‘lacking explanation’ and ‘compact’, 1.5% thought it was ‘comprehensive’ and ‘good’. **(b)** Report B was found ‘lengthy’ by 33.8% of the participants, 32.3% said it was ‘detailed’, 12.3% of the doctors said it was ‘exhaustive’, 6.2% found it ‘incomprehensible’, 4.6% said it was ‘easy’ and ‘useful’, 3.1% said the results were ‘noticeable’ and 1.5% said the format was ‘boring’ and ‘simple’.

### General Preference between Report A and Report B

The study group was questioned regarding their general preference for either of the two reports. We found 75.5% of the answers in favour of report A while 24.5% chose report B, as shown in Figure 5.

**Figure 5.**
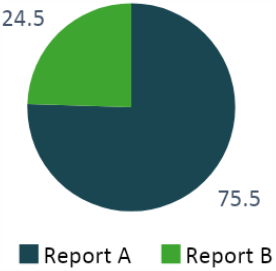
Report preference of the participants. 75.5% generally preferred report A while 24.5% chose report B.

With respect to the reason(s) for their selection, 38.8% of the participants choosing report A mentioned the ‘conciseness’ and 34.7% mentioned the ‘easier scientific and technical language’, while 8.2% preferred its ‘preciseness’. About 6.1% of the doctors commented on the ‘time efficiency’ of report A, while another 6.1% commented on its friendlier layout. Out of the 24.5% participants that preferred report B, 22.4% said it was more ‘detailed’, 4.1% liked the ‘Doctor friendly format’ and 2.0% found it ‘easy to interpret’ as shown in Figure 6.

**Figure 6.**
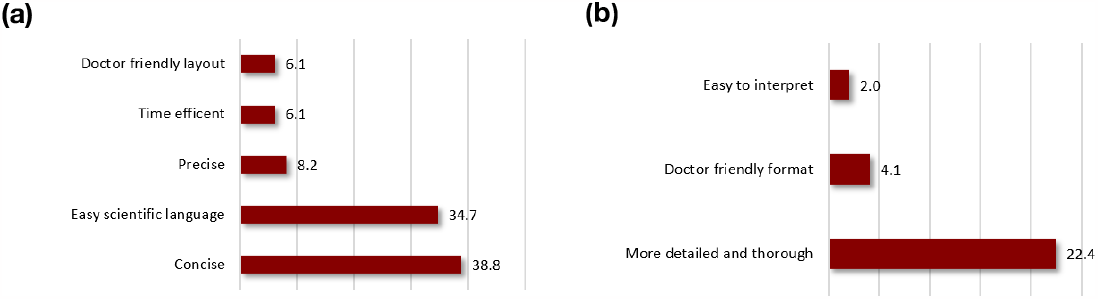
Reasons for preferring report A and report B: **(a)** 38.8% of those choosing report A mentioned the ‘conciseness’ and 34.7% mentioned the ‘easier scientific and technical language’, where 8.2% preferred its ‘preciseness’. About 6.1% of the doctors commented on the ‘‘time efficiency of report A, and its ‘friendlier layout’. **(b)** Out of the 24.5% participants that preferred report B, 22.4% said it was ‘more detailed’, 4.1% liked the ‘doctor friendly format’ and 2.0% found it ‘easy to interpret’.

### Ease of understanding of Report A and Report B

When asked about which report was easily comprehensible, 71.4% of the respondents chose report A (Figure 7). The respondents were also asked to explain their choice and the majority of them (47 out of the 49) did explain their answers.

**Figure 7.**
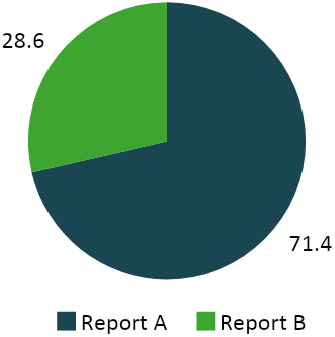
Report that’s easy to understand. 71.4% of participants found report A easier to understand whereas 28.6% found report B easier.

About 48.6% of the respondents found report A to be ‘concise and precise’, and 31.4% replied that it had an easier ‘language’, as shown in Figure 8a. About 14.3% preferred report A as they said it had a more ‘Doctor friendly format’ when compared to report B.

**Figure 8.**
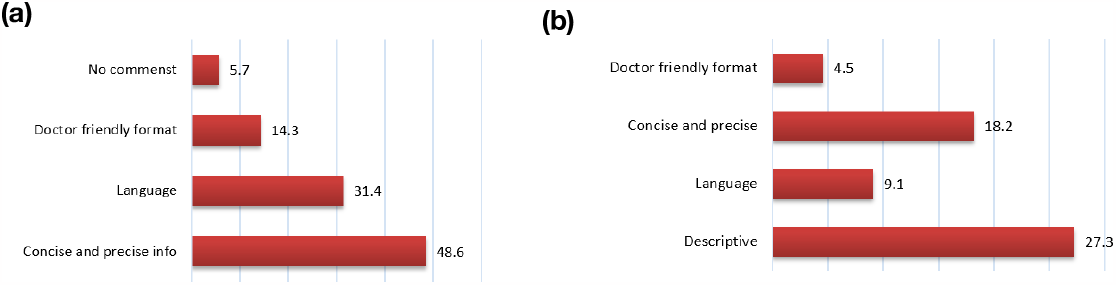
Reasons for ease of understanding of report A and B: **(a)** 48.6% of the participants found the information in report A to be ‘concise and precise’, 31.4% appreciated the ‘language’ used in report A, 14.3% found the format used to be ‘doctor friendly’ and 5.7% had ‘no comments’ regarding their preference. **(b)** 27.3% of the doctors found report B easier as it was ‘descriptive’, 9.1% favored the ‘language’ used, 18.5% found the report to be ‘concise and precise’ and 4.5% found the format to be ‘doctor friendly’.

About 27.3% of the doctors found report B easier as it was ‘descriptive’, 9.1% favoured the ‘language’ used, 18.2% found the report to be ‘concise and precise’ and 4.5% found the layout to be ‘doctor friendly’.

### Length of Reports A and Report B

According to the dominant majority (83.7%) of respondents, report A was of ‘appropriate length’ (Figure 9). But 16.3% reported to believe otherwise. Similarly 83.7% of the participants found report B lengthier than report A.

**Figure 9.**
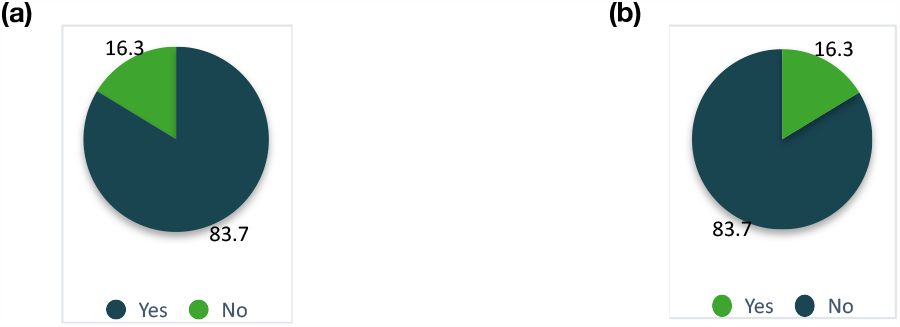
Appropriate length of both the reports: **(a)** About 83.7% of the participants found the length of report A to be appropriate whereas 16.3% disagreed. **(b)** 83.7% clinicians found report B to be lengthier than report A and 16.3 % found the length to be appropriate.

### Time taken to read through both the reports

According to 89.8% participants, Report B, which was a detailed genomic report, and was therefore more time consuming to read as compared to Report A (Figure 10).

**Figure 10.**
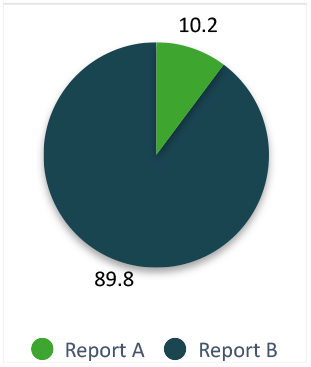
Time taken to read through both the reports: According to 89.8% participants report B was more time consuming to read compared to report A, while 10.2% responded otherwise.

### Difficulty of language used in both the reports

When asked about the difficulty of the language used in poet A, half of the respondents (51%) chose the word ‘moderate’. When asked about Report B, a third of the respondents (30.6%) used the word ‘Somewhat Hard’.

### Sections difficult to understand

Our survey also included a qualitative question on sections or parts of the report that were difficult to understand. A summary of the responses on the perceived difficulty in understanding the different sections of the two report templates is given in Figure 12. In Report A, 13.3% of the participants reported difficulty in comprehending the information provided under the ‘Carrier Risk’ section, while 4.4% found it challenging to understand the ‘Pharmacogenomics Association’ and the ‘Names of Genes/Alleles’. A smaller proportion of the respondents (2.2%) stated difficulty in comprehending the terms ‘Gene Transcripts’, ‘Summary’, and ‘Zygosity Variant’. However, a considerable majority of the participants (71.1%) did not respond to the question.

**Figure 11.**
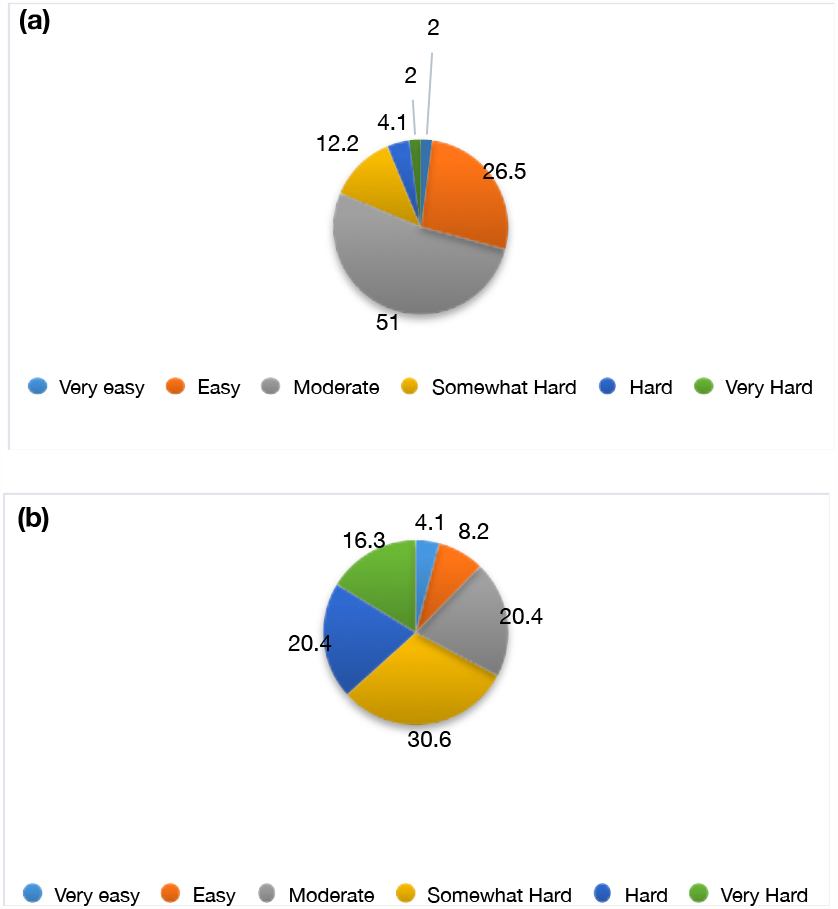
Difficulty level of the scientific and technical language of the reports: **(a)** 51% practitioners found the difficulty level of the language used in report A to be ‘moderate’, 26.5% found it ‘easy’, 12.2% found it ‘somewhat hard’ to understand, 4.1% found it ‘hard’ and 2% said it was ‘very hard’ and ‘very easy’. **(b)** About 30.6% clinicians found the level of difficulty to be ‘somewhat hard’, 20.4% said it was ‘moderate’ and ‘very easy’, 16.3% answered ‘very hard’, 8.2% found it ‘easy’ and 4.1% found it ‘very easy’ to understand.

**Figure 12.**
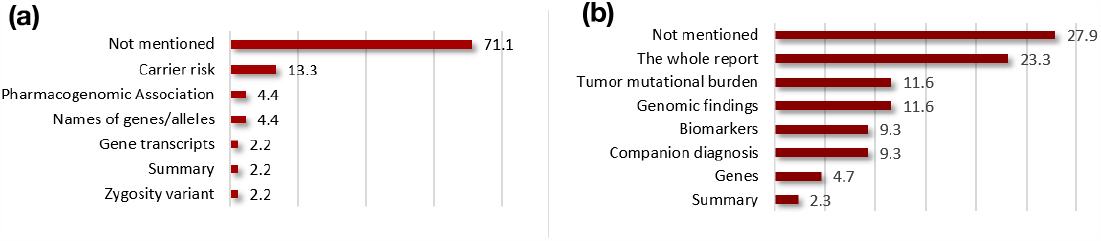
Sections difficult to understand in both the reports: **(a)** About 71.1% participants didn’t respond to the question. Meanwhile 13.3% couldn’t understand ‘carrier risk’ and 4.4% said they had difficulty with the terms ‘pharmacogenomics association’ and the ‘names of genes/alleles’, 2.2% stated that they were unable to understand ‘gene transcripts’, ‘summary’ and ‘zygosity variant’. **(b)** In report B, 27.9% participants chose not to answer the question, while 23.3% said they were unable to comprehend the whole report. About 11.6% said they couldn’t understand what ‘tumor mutational burden’ and ‘genomic findings’ meant, 9.3% mentioned difficulty comprehending ‘biomarkers’ and ‘companion diagnosis’, 4.7% didn’t understand the various mentioned ‘genes’ section and 2.3% had difficulty understanding the ‘summary’ section in report B.

Conversely, in Report B, 23.3% of the participants expressed difficulty in understanding the entire report, while 11.6% found the terms ‘Tumor Mutational Burden’ and ‘Genomic Findings’ challenging to comprehend. About 9.3% of the clinicians reported difficulty in grasping the terms ‘Biomarkers’ and ‘Companion Diagnosis’. Similar to the case with Report A, although to a lesser extent, the majority of the participants (27.9%) chose not to answer the question.

### Actionability

When clinicians were questioned about their confidence and therefore ability to carry out clinical decisions regarding any treatment intervention, based on the information given in both reports, 73.5% answered positively that they would be able to, as shown in Figure 13.

**Figure 13.**
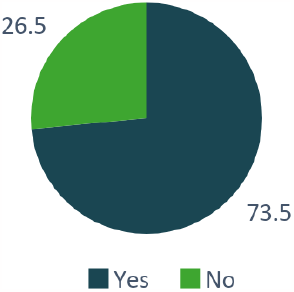
Would you be able to take action? 73.5% participants were confident that they would be able to take further actions based on the information present in the reports whereas 26.5% answered that they would

When asked for the reasons behind their confidence, the dominant majority chose not to answer the question. However, some participants stated the reasons for their ability to take action. For example, amongst those who responded positively when asked about their ability to take action, 2.8% said they would be able to proceed with an intervention but only ‘with the information provided in report A’, another 2.8% assured that they might only ‘identify risk factors and the genes’ (Figure 14a). Participants who responded negatively to the question on their ability to make action, 15.4% considered the evidence in the reports to be insufficient, and 7.7% said they will be capable of carrying out treatment management only ‘if report A was more detailed’ as shown in Figure 14b whilst the other 7.7% stated that they were only capable of ‘referral of the patient’ to a specialist, based on the information provided in the reports.

**Figure 14.**
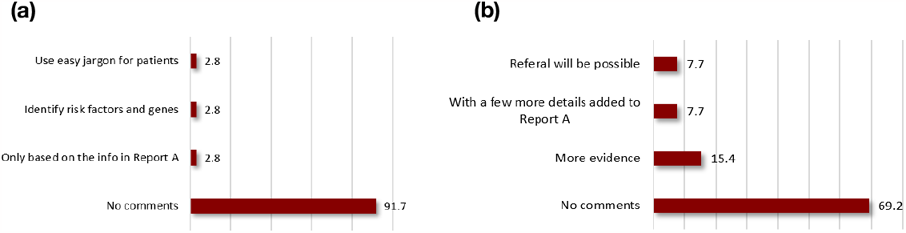
Ability to take action based on the reports. **(a)** Those who said ‘yes’: Based on the data given in the reports, 2.8% of the doctors said they will be able to action based on the ‘information given in report A’, they will only be able to ‘identify the risk factors and genes’ and if there is ‘easy jargon used for patients’ to be able to understand the report as well. An overwhelming majority (91.7%) refrained from commenting. **(b)** Those who said ‘no’: Inability to take action was based on 15.4% saying that there was ‘lack of evidence’,7.7% said only if ‘report A had more details’ and only ‘referral will be possible’, while 69.2% didn’t comment.

### Perceived reliability of the reports

Finally, the clinicians were also questioned on how reliable did they think the information in the both the genomic reports was; 87.8% answered that they found the reports reliable, while the remaining 12.2% were skeptical of the given information, as shown in Figure 15.

**Figure 15.**
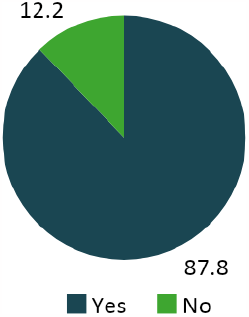
Reliability of the reports. About 87.8% participants found the reports reliable whereas 12.2% were not satisfied.

Both groups of participants were asked for the reasons behind the reliability of these reports or lack thereof. Out of the majority group which found the information in the reports reliable, most did not present an answer, but 2.3% showed a tilt in their confidence toward Report B as they believed it contained ‘more clinical evidence’ and anther 2.8% presumed that ‘a doctor would only order tests from a trustworthy lab’ (Figure 16a).

**Figure 16.**
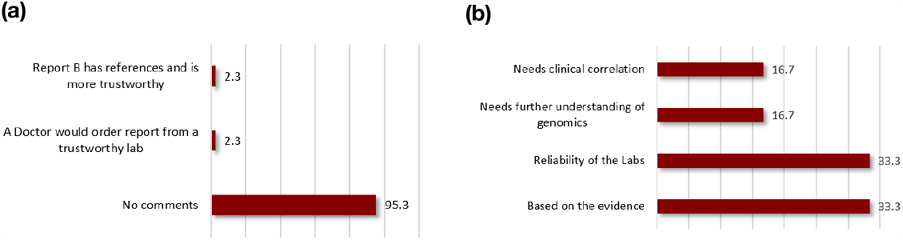
Reliability of the genomic reports. **(a)** From the majority group which was positive about the reliability of the reports, 2.3% trust the choice of their labs and another 2.3% believed that report B is more detailed and therefore more trustworthy’ but an overwhelming majority (95.3%) respondents did not answer this question. **(b)** From the group which was skeptical of the reliability of the two reports, around 33.3% cited the ‘lack of evidence’ and another 33.3% citing the ‘reliability of the labs’ as their reasons. Around 16.7% mentioned the need for more genomic literacy and another 16.7% mentioned the ‘need for clinical correlation’.

From the group which was skeptical with regards to the reliability of the two reports, around 33.3% cited that the reports ‘lacked sufficient evidence’ while another 33.3% were unconvinced with the ‘reliability of the laboratories’. Around 16.7% of the clinicians cited their ‘need of further understanding in genomics’ and another 16.7% stated that the reports needed ‘more clinical correlation’ with the diagnosis mentioned in the reports.

### Recommendations for designing better genomics test reports

The survey ended with a subjective and qualitative question asking each respondent for ‘Suggestions that you would like to give, in order to make genomics test report easy for doctors to understand’. Around half the participants responded with an answer to this question. When asked for their recommendations, 12.2% of participants preferred a genomic report to be ideally short and concise, whereas 10.2% suggested a non-specialist, friendly format and 4.1% recommended a tabulated format for reporting of genomic test results. The complete list of suggestions is summarised in Figure 17.

**Figure 17.**
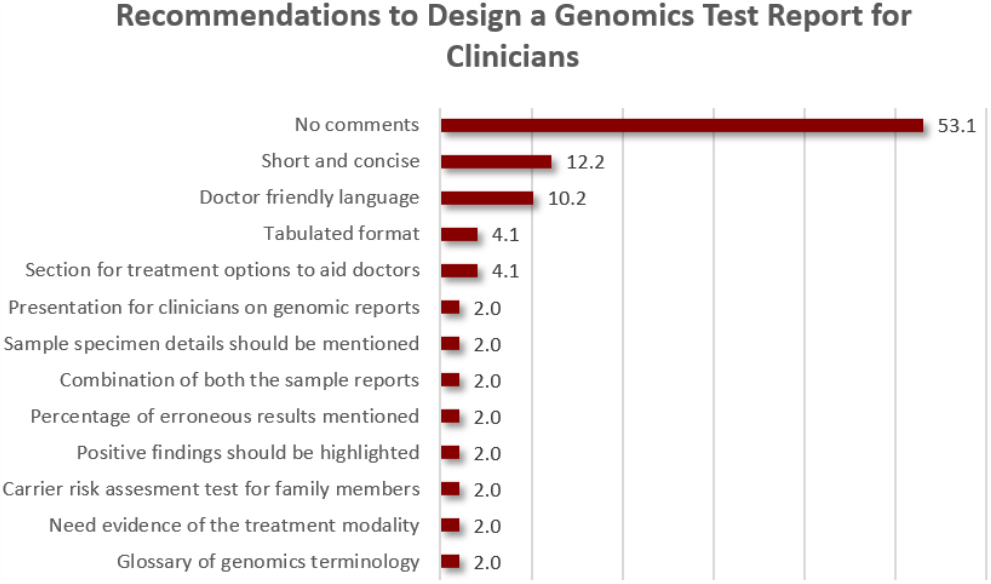
Recommendations for designing genomic reports for clinicians. About 12.2% of the clinicians opted for ‘short and concise’ reports, 10.2% suggested the use of ‘doctor friendly language’ and 4.1% would prefer a ‘tabulated layout’ and a separate ‘section for treatment options to help doctors’. 2.0% of the participants suggested the following: ‘presentations for clinicians on genomic reports’, mentioning the ‘specimen details’, ‘combination of both report A and B’ would be ideal, ‘percentage of erroneous results mentioned’, ‘positive findings should be highlighted’, ‘carrier risk assessment test’ should be conducted for families, ‘more evidence for treatment modalities’ and a ‘glossary for genomics terminology’ should be added at the end of the report.

We also went on and condensed these 23 individual suggestions given by the participants into a total of 11 recommendations and classified them into 4 categories: 1) Language, 2) Layout of the report, 3) content and 4) Reliability and actionability. These 11 recommendations are laid down in Table 1.

**Table 1.** Participants were asked for ‘Suggestions that you would like to give, in order to make genomics test reports easy to understand’ and a total of 23 suggestions were collected from the participants that were grouped into four categories of 11 final recommendations. The categories were: a) Language b) Content c) Layout and d) Reliability of the genomic reports.

| Category | Recommendations | Detail |
| --- | --- | --- |
| Language | • Use minimal jargon<br>• Keep it short and concise | • Try to use language which is understood by all specialties, even general physicians and dentists.<br>• Avoid complicated details and make the report short and easy to understand. |
| Content | • Specimen details should be more prominent.<br>• Provide a glossary of all genetic and genomic terms.<br>• Add a section of treatment options<br>• Add a Carrier Risk Assessment test recommendation for family members, if any.<br>• Combination of both report A and B. | • Patient details and specimen information should be added on every page.<br>• A glossary of terms and technical details would help both clinician and patient to understand the reports better. |
| Layout | • Use a tabulated format.<br>• Positive findings should be highlighted. | • The tabulated layout was preferred for ease of reading.<br>• A good layout can enhance better comprehension, reduce time and stress and improve the efficiency of the doctor. |
| Reliability and Actionability | • Build and nurture trust in laboratories that are offering genomic tests.<br>• Add more information on clinical correlation of the results. | • Avoid overuse of graphics and logos unless real required, e.g. to prove authenticity of the lab.<br>• Try to add latest clinical references relevant to the report findings, in order to strengthen the confidence in the diagnosis, and provide a separate section on what to do next. |

## Discussion

Genomic testing is becoming increasingly integrated into standard testing protocols throughout the world, indicating that clinicians are likely to order these tests as part of routine patient care. As a result, it is essential that genomic test reports convey the results and their significance in a clear and concise manner to clinicians without a genomics background. Our study indicates a pressing need for an well-designed and user-tested genomic report template that is accessible to both medical specialists and non-specialist practitioners in Pakistan.

The study used a survey instrument to seek input from clinicians at different stages of their careers who are working around the Precision Medicine Lab and the Centre for Genomic Sciences in Peshawar, Pakistan and analysed a total of 49 survey responses, wherein participants evaluated two different report templates, that are widely used especially in the North American context. Report A was favoured by a majority of clinicians (75.5%) due to its conciseness, as indicated by 38.8% of participants, and its use of simpler scientific language, as stated by 34.7% of respondents. When asked to indicate ‘Which report was easy to understand?’ The results revealed a clear preference for report A among the majority of the practitioners (71.4%). Additionally, 31.4% of the respondents favoured report A due to its use of straightforward language and 14.3% preferred the doctor-friendly format.

During the survey, the participants were also asked for their views on the appropriate length of a genomic test report (between the two given templates) and the results indicated that 83.7% of the respondents preferred report A over report B. Furthermore, 89.8% of the participants perceived report B to be more time-consuming to comprehend, which could be attributed to its length and scientific language used. Notably, 20.4% of the clinicians reported that the language in report B was ‘hard’, and 16.3% stated it was ‘very hard’, whereas only 4.1% found the language in report B to be ‘very easy’.

When questioned regarding their ability to undertake action based on the reports, a considerable proportion of the participants (73.5%) agreed that they could were comfortable to proceed with patient management based on the genomic test results. However, when asked for the rationale behind their actionability, 91.7% of the participants refrained from providing any explanation. Similarly, 69.2% of the participants chose not to respond to queries about their inability to take action, while 15.4% of the participants stated that the information presented required further clinical evidence before they could proceed with patient treatment.

The participants were also asked about the reliability of the genomic report templates, and 87.8% of them responded that they found the reports reliable. In contrast, 33.3% of the respondents who considered the reports unreliable expressed doubts about the sufficiency of the evidence regarding the diagnosis in the reports, especially with respect to variants that are specific to our region. During focus group discussions, Cutting *et al* observed that clinicians expressed a desire for references to academic literature that support the interpretations presented in the report, which is consistent with the concern expressed by the participants of our study as well (Cutting *et al*., 2016). However, a majority of the participants (95.3%) chose not to provide any justification for their answers.

This non-responsive pattern may be indicative of a lack of full understanding of genomics-related concepts, the specialised language or both (Barakzai *et al*, 2022). This could partially supported by the difficulty encountered and self-reported by participants of this study in comprehending several critical sections in both reports, which are integral to delivering comprehensive patient care. This potentially indicates a lack of genomic literacy among the healthcare professionals in the country as mentioned by (Riaz *et al*., 2019). This could be pointing at a need for additional training in understand genomic technologies and diagnostic results that are offered by such methods in the lab.

Approximately 79.6% of our survey respondents were non-specialists, comprising of trainee medical officers, MPhil students, and medical officers, which may lead to potential bias in our study with more respondents who might not yet have been exposed to diagnosis, prognosis and treatment based on genomic tests. Nonetheless, this underscores the pressing need to bridge the gap in genomic literacy by designing reports that are more accessible to healthcare providers across the board. This can potentially help in increasing the adoption of genetic testing in the clinic and therefore in enabling the medical fraternity to provide more efficacious and personalised treatments to patients, ultimately improving patient outcomes.

The study findings confirm several conclusions drawn from prior research, particularly the importance of developing a user-friendly genomic report (Haga *et al*., 2014) as indicated in Figure 14a where the study participants suggested that the reports should use ‘easy jargon for patients’ so that not only are they comprehensible to the clinician, but can also be understood by the patients as well (Stuckey *et al*., 2015; Williams *et al*., 2016; Joseph *et al*., 2017). With most previous studies focused on the needs of the patient as at the user, we focus on the clinician being the primary user and the key decision maker in the clinical setting especially in the Pakistani context.

Moreover, participants’ responses highlighted the need for effective communication of potential risks to the patient, as noted by Lautenbach *et al (*Lautenbach *et al*, 2013). Our recommendations also align with the need to include technical information and limitations that are raised by previous works (Dorschner *et al*., 2014; Kelman *et al*., 2016).

This study emphasises the need for a framework for the effective reporting of a genomic test result and its effective communication in an accessible manner to the doctor, that is contextualised in the local practice. Implementing the recommendations outlined in this study can help us begin to enable genomic reports to clearly communicate results to doctors, and ideally the patients, without requiring the assistance of a genomics expert or other medical specialists. But we do understand, that more iterations with the users of these reports (doctors and patients) will further improve the reporting and the overall communication. It is also a given, that one cannot rule out the need for consulting the experts in some cases for further discussion and treatment planning. In summary, a clear and comprehensive explanation of the test results, along with guidance on next steps, can help doctors and patients make informed decisions about their healthcare along with the help of their healthcare practitioner.

Like every study, this survey has its sampling bias, i.e. the over-representation of respondents from one geography, as the majority of participants were located in Peshawar, Pakistan, and the over-representation of respondents trained in dentistry (BDS; 63.3%) compared to medicine and surgery (MBBS; 36.7%) and the overwhelming majority of non-specialists (79.6%) to specialists (20.2%). This uneven distribution of participants may have exerted a discernible, and perhaps avoidable, impact on the outcomes of our study, particularly with respect to the comprehension of details on both genomic test reports that were tested in this study. This warrants for a larger and more even sample size in any subsequent research.

## Conclusion

Our study highlights the importance of a user-centred approach in the design of genomic reports. The user in a Pakistani clinical context is the clinician who is the primary decision maker for various reported reasons. By obtaining feedback from clinicians from different backgrounds, from all levels of training or practice and from different institutions, we have developed recommendations that can be applied to the reporting genomic test reports, enabling better-informed decision making. The findings of this study have important implications for the future of genomic report design for clinicians in Pakistan, and we hope they will contribute to the development of more user-friendly and accessible reports that can help in increasing the adoption of genomic testing in the clinic and ultimately help in improving patient outcomes.

## Data Availability

All data produced in the present study are available upon reasonable request to the authors.

## Notes

### Competing Interest Statement

The authors have declared no competing interest.

### Funding Statement

This study was supported by Precision Medicine Lab, a federally-funded project under the National Centre of Big Data and Cloud Computing (NCBC) by the Higher Education Commission Pakistan.

## References

Afroze, B., Humayun, K. N. and Qadir, M. (2008) ‘Newborn screening in Pakistan - lessons from a hospital-based congenital hypothyroidism screening programme.’, Annals of the Academy of Medicine, Singapore, 37(12 Suppl), pp. 113–114.

Ashfaq, M. et al. (2013) ‘The views of Pakistani doctors regarding genetic counseling services - is there a future?’, Journal of genetic counseling, 22(6), pp. 721–732. doi: 10.1007/s10897-013-9578-2.

Barakzai, M., Rehmat, S. and Khan, F. (2022) ‘Abstract 6337: Assessing genomic literacy among medical trainees and practitioners in Pakistan’, Cancer Research, 82, p. 6337. Available at: 10.1158/1538-7445.AM2022-6337.

Brett Gemma R., et al. “Co-design, implementation, and evaluation of plain language genomic test reports.” NPJ Genomic Medicine 7.1 (2022): 61

Cresswell, L. et al. (2020) ‘General Genetic Laboratory Reporting Recommendations’, Association for Clinical Genetic Science, pp. 1–11.

Cutting, E. et al. (2016) ‘User-centered design of multi-gene sequencing panel reports for clinicians’, Journal of Biomedical Informatics, 63, pp. 1–10. Available at: 10.1016/J.JBI.2016.07.014.

Dang, B. N. et al. (2013) ‘Examining the Link between Patient Satisfaction and Adherence to HIV Care: A Structural Equation Model’, PLoS ONE, 8(1), pp. 1–10. doi: 10.1371/journal.pone.0054729.

Dimatteo, M. R. (2004) ‘The role of effective communication with children and their families in fostering adherence to pediatric regimens.’, Patient education and counseling, 55(3), pp. 339–344. doi: 10.1016/j.pec.2003.04.003.

Dorschner, M. O. et al. (2014) ‘Refining the structure and content of clinical genomic reports.’, American journal of medical genetics. Part C, Seminars in medical genetics, 166C(1), pp. 85–92. doi: 10.1002/ajmg.c.31395.

Elder, N. C. and Barney, K. (2012) ‘“But what does it mean for me?” Primary care patients’ communication preferences for test results notification.’, Joint Commission journal on quality and patient safety, 38(4), pp. 168–176. doi: 10.1016/s1553-7250(12)38022-7.

Farmer, G. D. et al. (2020) ‘Recommendations for designing genetic test reports to be understood by patients and non-specialists.’, European journal of human genetics : EJHG, 28(7), pp. 885–895. doi: 10.1038/s41431-020-0579-y.

Furqan, A. (2021) Establishing the Pakistani Society of Medical Genetics. Available at: https://perspectives.nsgc.org/Article/establishing-the-pakistani-society-of-medical-genetics (Accessed: 24 October 2021).

Haga, S. B. et al. (2014) ‘Developing patient-friendly genetic and genomic test reports: formats to promote patient engagement and understanding.’, Genome medicine, 6(7), p. 58. doi: 10.1186/s13073-014-0058-6.

Heggie, J. (2019) ‘Genomics: a revolution in health care?’, National Geographic. Available at: https://www.nationalgeographic.com/science/article/partner-content-genomics-health-care.

Joseph, G. et al. (2017) ‘Information Mismatch: Cancer Risk Counseling with Diverse Underserved Patients.’, Journal of genetic counseling, 26(5), pp. 1090–1104. doi: 10.1007/s10897-017-0089-4.

Kelman, A. et al. (2016) ‘Communicating laboratory test results for rheumatoid factor: What do patients and physicians want?’, Patient Preference and Adherence, 10, pp. 2501–2517. doi: 10.2147/PPA.S104396.

Khoury, M. J., Burke, W. and Thomson, E. (2009) ‘Genetics and Public Health in the 21st Century: Using Genetic Information to Improve Health and Prevent Disease’, Genetics and Public Health in the 21st Century: Using Genetic Information to Improve Health and Prevent Disease, pp. 1–660. doi: 10.1093/acprof:oso/9780195128307.001.0001.

Lautenbach, D. M. et al. (2013) ‘Communicating genetic risk information for common disorders in the era of genomic medicine.’, Annual review of genomics and human genetics, 14, pp. 491–513. doi: 10.1146/annurev-genom-092010-110722.

Linn, A. J. et al. (2013) ‘May you never forget what is worth remembering: the relation between recall of medical information and medication adherence in patients with inflammatory bowel disease.’, Journal of Crohn’s & colitis, 7(11), pp. e543–50. doi: 10.1016/j.crohns.2013.04.001.

Martin, L. R. et al. (2005) ‘The challenge of patient adherence.’, Therapeutics and clinical risk management, 1(3), pp. 189–199.

Matthijs, G. et al. (2016) ‘Guidelines for diagnostic next-generation sequencing.’, European journal of human genetics : EJHG, 24(1), pp. 2–5. doi: 10.1038/ejhg.2015.226.

McGovern, M. M., Benach, M. and Zinberg, R. (2003) ‘Interaction of genetic counselors with molecular genetic testing laboratories: implications for non-geneticist health care providers.’, American journal of medical genetics. Part A, 119A(3), pp. 297–301. doi: 10.1002/ajmg.a.20196.

Miller, D. T. et al. (2010) ‘Consensus statement: chromosomal microarray is a first-tier clinical diagnostic test for individuals with developmental disabilities or congenital anomalies.’, American journal of human genetics, 86(5), pp. 749–764. doi: 10.1016/j.ajhg.2010.04.006.

Ostergren, J. E. et al. (2015) ‘How Well Do Customers of Direct-to-Consumer Personal Genomic Testing Services Comprehend Genetic Test Results? Findings from the Impact of Personal Genomics Study.’, Public health genomics, 18(4), pp. 216–224. doi: 10.1159/000431250.

Recchia, G. et al. (2020) ‘Creating genetic reports that are understood by nonspecialists:a case study’, Genetics in Medicine, 22(2), pp. 353–361. doi: 10.1038/s41436-019-0649-0.

Riaz, M. et al. (2019) ‘Implementation of public health genomics in Pakistan’, European Journal of Human Genetics, 27(10), pp. 1485–1492. doi: 10.1038/s41431-019-0428-z.

Richards, S. et al. (2015) ‘Standards and guidelines for the interpretation of sequence variants: a joint consensus recommendation of the American College of Medical Genetics and Genomics and the Association for Molecular Pathology.’, Genetics in medicine : official journal of the American College of Medical Genetics, 17(5), pp. 405–424. doi: 10.1038/gim.2015.30.

Sandhaus, L. M. et al. (2001) ‘Reporting BRCA test results to primary care physicians.’, Genetics in medicine : official journal of the American College of Medical Genetics, 3(5), pp. 327–334. doi: 10.1097/00125817-200109000-00001.

Shaer, O. et al. (2015) ‘Informing the Design of Direct-to-Consumer Interactive Personal Genomics Reports’, Journal of Medical Internet Research, 17(6), p. e146. Available at: 10.2196/jmir.4415.

Sirisena, N. et al. (2016) ‘The Provision of Medical and Health Genetics and Genomics in the Developing World’, Medical and Health Genomics, pp. 285–294. doi: 10.1016/B978-0-12-420196-5.00021-6.

Stuckey, H. et al. (2015) ‘Enhancing genomic laboratory reports from the patients’ view: A qualitative analysis.’, American journal of medical genetics. Part A, 167A(10), pp. 2238–2243. doi: 10.1002/ajmg.a.37174.

Weingarten, S. R. et al. (1995) ‘A study of patient satisfaction and adherence to preventive care practice guidelines.’, The American journal of medicine, 99(6), pp. 590–596. doi: 10.1016/s0002-9343(99)80243-5.

Williams, J. L. et al. (2016) ‘Enhancing genomic laboratory reports: A qualitative analysis of provider review.’, American journal of medical genetics. Part A, 170A(5), pp. 1134–1141. doi: 10.1002/ajmg.a.37573.

